# Blood-based screening for HPV-associated cancers

**DOI:** 10.1101/2024.01.04.24300841

**Authors:** Dipon Das, Shun Hirayama, Ling Aye, Michael E. Bryan, Saskia Naegele, Brian Zhao, Vasileios Efthymiou, Julia Mendel, Adam S. Fisch, Lea Kröller, Birgitta E. Michels, Tim Waterboer, Jeremy D. Richmon, Viktor Adalsteinsson, Michael S. Lawrence, Matthew G. Crowson, A. John Iafrate, Daniel L. Faden

**Affiliations:** Department of Otolaryngology-Head and Neck Surgery, Harvard Medical School, Boston, Massachusetts; Massachusetts Eye and Ear, Boston, Massachusetts; Department of Otorhinolaryngology-Head and Neck Surgery, Wakayama Medical University, Wakayama, Japan; Department of Pathology, Massachusetts General Hospital, Boston, Massachusetts; Division of Infections and Cancer Epidemiology, Deutsches Krebsforschungszentrum, Heidelberg, Germany; Broad Institute of MIT and Harvard, Cambridge, Massachusetts; Krantz Family Center for Cancer Research, Massachusetts General Hospital Cancer Center, Boston, Massachusetts

**Author notes:** Corresponding Author, Corresponding author: Daniel L. Faden, MD, Department of Otolaryngology-Head and Neck Surgery, Harvard Medical School. Division of Head and Neck Surgical Oncology, Massachusetts Eye and Ear 243 Charles St, Boston, MA 02118. Equal contribution (Shun Hirayama, Ling Aye, Michael E. Bryan, Saskia Naegele).

## Abstract

**Background:** HPV-associated oropharyngeal cancer (HPV+OPSCC) is the most common HPV-associated cancer in the United States yet unlike cervical cancer lacks a screening test. HPV+OPSCCs are presumed to start developing 10-15 years prior to clinical diagnosis. Circulating tumor HPV DNA (ctHPVDNA) is a sensitive and specific biomarker for HPV+OPSCC. Taken together, blood-based screening for HPV+OPSCC may be feasible years prior to diagnosis.

**Methods:** We developed an HPV whole genome sequencing assay, HPV-DeepSeek, with 99% sensitivity and specificity at clinical diagnosis. 28 plasma samples from HPV+OPSCC patients collected 1.3-10.8 years prior to diagnosis along with 1:1 age and gender-matched controls were run on HPV-DeepSeek and an HPV serology assay.

**Results:** 22/28 (79%) of cases and 0/28 controls screened positive for HPV+OPSCC with 100% detection within four years of diagnosis and a maximum lead time of 7.8 years. We next applied a machine learning model classifying 27/28 cases (96%) with 100% detection within 10 years. Plasma-based PIK3CA gene mutations, viral genome integration events and HPV serology were used to orthogonally validate cancer detection with 68% (19/28) of the cohort having multiple cancer signals detected. Molecular fingerprinting of HPV genomes was performed across patients demonstrating that each viral genome was unique, ruling out contamination. In patients with tumor blocks from diagnosis (15/28), molecular fingerprinting was performed within patients confirming the same viral genome across time.

**Conclusions:** We demonstrate accurate blood-based detection of HPV-associated cancers with lead times up to 10 years before clinical cancer diagnosis and in doing so, highlight the enormous potential of ctDNA-based cancer screening.

## Introduction

Circulating tumor (ct) DNA approaches are being applied across nearly all cancer care contexts, including screening. Numerous Multi-Cancer Early Detection (MCED) approaches are now available in both research and clinical settings^1–3^. Continued advancements in the platforms and computational approaches available are driving lower and lower limits of cancer detection in the blood. How early in development a cancer can be accurately detected remains to be determined^4^.

ctDNA-based early detection approaches have the potential to drastically improve cancer screening, particularly for cancers that currently do not have screening approaches. At present, only four cancer types have population-level screening tests recommended, meaning that most diagnoses are in cancer types for which there is no screening and thus tend to be later-stage disease, when the cancer has reached a size to become symptomatic^5,6^. Because diagnosis of later-stage cancers results in decreased survival, increased costs, and increased treatment morbidity, detection of asymptomatic, early-stage cancers creates a unique opportunity for simultaneous improvement in all three of these critical domains^7–12^.

HPV-associated oropharyngeal squamous cell carcinoma (HPV+OPSCC) is the most common HPV- associated malignancy in the United States, yet unlike HPV-associated cervical cancer, it lacks a screening test^13,14^. Oropharyngeal infection with HPV typically occurs >20 years prior to clinical cancer diagnosis and genomic models suggest HPV+OPSCC begins developing 10-15 years prior to diagnosis^15^. Further, patients with HPV+OPSCC develop HPV oncoprotein-directed antibodies that have high sensitivity and specificity for HPV+OPSCC at the time of diagnosis and can be detected >10 years before diagnosis, in some patients^16–19^. HPV+OPSCCs develop inside the deep crypts of the palatine and lingual tonsils and thus are frequently asymptomatic, even at the time of clinical diagnosis. Instead, the most common presenting symptom for HPV+OPSCC is a palpable neck mass, as HPV+OPSCCs metastasize to cervical lymph nodes in nearly all cases by the time of diagnosis. Taken together, there is a substantial time window in which screening early detection of HPV+OPSCC may be feasible and enable detection of earlier-stage, non-metastatic cancers.

HPV+OPSCCs release fragments of the viral cancer genome, termed circulating tumor HPV DNA (ctHPVDNA), which is detectable at diagnosis with high sensitivity and specificity and is more reliable than mutation- or methylation-based ctDNA detection^20,21^. Importantly, while transient oropharyngeal infection with HPV is common, it does not lead to detectable ctHPVDNA^22^. A small proof-of-principle study has suggested that ctHPVDNA may be detectable in asymptomatic individuals years prior to clinical diagnosis of HPV+OPSCC, but that existing approaches are not sensitive enough for reliable ctHPVDNA detection in most cases^23,24^.

In this retrospective study, we evaluate the feasibility of a screening test for HPV-associated cancers in asymptomatic persons, aiming to determine if a multi-modal next-generation sequencing (NGS) liquid biopsy can reliably detect ctHPVDNA and how far before clinical diagnosis this detection can occur.

## Methods

### Participants

#### Diagnostic cohort

5-10ml of whole blood was collected from 153 patients presenting with HPV+OPSCC at Mass Eye and Ear in Boston, Massachusetts. HPV+OPSCC diagnosis consisted of histopathologic confirmation of SCC, p16 immunohistochemistry, and direct HPV testing with RNA in-situ hybridization or DNA polymerase chain reaction (PCR). 5-10ml of whole blood was also collected from 153 patients presenting for routine care at Mass Eye and Ear– 100 patients had no history of cancer, and 53 patients had a history of non-HPV-associated cancers.

#### Screening cohort

The MassGeneralBrigham biobank has prospectively collected blood samples and data from 140,000 patients presenting for care in the healthcare system based in Boston, Massachusetts. Biobank clinical data was queried to identify patients who: 1. were diagnosed with HPV+OPSCC >1 year after blood sample contribution, 2. had ≥1ml of plasma available for retrieval and, 3. had no history of an HPV-associated malignancy at the time of blood sample contribution. Medical record review was then undertaken to ensure confirmation of HPV+OPSCC development and extraction of clinicodemographic data, yielding a cohort of 28 patients with 1-4ml of frozen plasma. 20/28 patients were eventually diagnosed within MassGeneralBrigham permitting retrieval of fresh-frozen paraffin-embedded (FFPE) tumor tissue blocks from the time of diagnosis in 15 patients. 1:1 age- and sex-matched controls were identified in the biobank and matching plasma volumes were obtained.

### Sample processing

Cell free (cf) DNA was extracted from 1-5ml of frozen plasma as previously described for all cases and controls^20^. DNA was extracted from FFPE blocks after tumor identification and microdissection and sheared prior to sequencing. A custom hybrid capture library was designed with 1x end-to-end tiling over the full HPV genome of 43 genotypes (Supplementary Table S1). Libraries were prepared following the KAPA HyperCap cfDNA Workflow v1.0 (Roche). The steps include: 1. End Repair & A-Tailing; 2. KAPA Universal UMI Adapter Ligation; 3. Post-Ligation Cleanup; 4. Amplification using KAPA UDI primer mixes; 5. Cleanup using KAPA HyperPure Beads; 6. Quantification by Qubit and quality check by Tapestation; 7. Preparation of 4-plex DNA sample library pools; 8. Use of customized KAPA HyperCap Target Enrichment Probes for hybridization and amplification of the target libraries; 9. Cleanup of enriched target libraries using KAPA HyperCapture Bead Kit; 10. Quantification and quality check by Qubit and Tapestation; 11. Pooling and normalization of libraries; 12. Denaturation; and 13. Paired-end sequencing on an Illumina platform to ∼6000x coverage per sample. Optimal thresholds for sensitivity and specificity were established using serial dilutions (1ng – 0.00001ng) of HPV+OPSCC patient cfDNA into a mixture of control patient cfDNA run in triplicate (Supplementary Table S2).

### Sample analysis

Sequencing reads underwent adapter trimming and quality filtering prior to alignment by BWA-MEM to a combined reference genome encompassing human hg38 and 64 HPV reference genomes. After Unique Molecular Index (UMI) identification and deduplication, HPV genotypes were inferred from mapping to an HPV reference genome. The number of unique HPV-mapped UMIs in each sample was used to determine the ctHPVDNA read counts. For HPV16 samples with > 50% genome coverage, position-wise base counts were obtained, and a major haplotype was created by taking the most prevalent nucleotide at each position. These haplotypes were fed into the multi-alignment tool MUSCLE, along with multiple reference builds for HPV16 sublineages. The resulting alignment was fed into PhyML (run using TOPALi v2.5), assuming a transversion model, allowing for invariant sites and specifying a gamma distribution. The resulting phylogenetic tree enabled sublineage assignment of each sample by nearest distance to a sublineage reference genome.

For each sample, the HPV genotype and sublineage were inferred, and the data was re-aligned to the appropriate sublineage reference genome. Clonal mutations were called using bcftools, filtered based on allele fraction (> 0.5) and read depth (≥4), and then manually reviewed in Integrated Genomics Viewer (IGV). For cross-sample contamination analysis, samples were considered unique if they harbored a pattern of variants not present in any other sample. For tumor-plasma sample concordance analysis, the mutation patterns of plasma and tumor samples from the same patient were compared, to ensure consistency. Viral integration events were identified from viral-human chimeric reads after alignment to the combined HPV and human reference genome, with high confidence assigned to events having at least two supporting reads and HPV coverage between 10% and 95% of the entire contig. Single-nucleotide variants (SNVs) in PIK3CA were called using MuTect v1 and annotated by SnpEff. All mutation calls were manually reviewed in IGV by three researchers independently (LA, SH, DD) to remove low-evidence mutations and artifacts.

### Machine learning

A binary machine learning task was employed to classify samples into HPV-associated cancer or no cancer. Before modeling, standard data preprocessing techniques were implemented. Min-max normalization was performed on numeric features. We allocated 80% of the data for training and held out 20% for testing. We trained several interpretable model architectures, including Random Forest, AdaBoost, and Naive Bayes. Model performance metrics included accuracy, precision, recall, macro F1- score, specificity, negative predictive value (NPV), and the area under the receiver operating characteristic curve (AUC-ROC). Hyperparameter tuning was conducted using 10-fold cross-validation on the training set for each model. We computed feature importance for the interpretable tree-based models to identify influential predictors. Once the models were tuned, they were finalized, and bootstrap resampling was conducted on the held-out test set to generate empirical 95% confidence intervals for the models’ performance metrics. AUC-ROC curves were generated for the cross-validation and test sets using Stratified K-Fold cross-validation with five splits. To interpret the models, we utilized the permutation importance method. This method was applied to calculate and normalize the feature importance scores for all models. We visualized the feature importance to evaluate the impact of each feature on the models’ predictive performance.

### HPV serology

Multiplex serology testing for HPV 16, 33, 35, and 45 was performed at the German Cancer Research Center (DKFZ) as previously described^25^.

### Study oversight

The MassGeneralBrigham Institutional Review Board approved this study. Patients underwent informed consent. All the authors collected and analyzed the data, contributed to the drafting of the manuscript, and vouch for the completeness and accuracy of the data, the accuracy of the analyses, and the fidelity of the study to the protocol. No one who is not an author contributed to the manuscript.

## Results

### HPV-DeepSeek performance and training

Following assay design, optimization, and threshold setting, cfDNA from 153 patients presenting with HPV+OPSCC and 153 general adult population control patients were run on HPV-DeepSeek (sensitivity 98.7%, specificity 98.7%) (Figure 1A, B, C). HPV-DeepSeek data output was then used to train and test machine learning models for classifying HPV+OPSCC vs no HPV+OPSCC. The machine learning models had equivalent performance with overlapping confidence intervals (Supplementary Table S3). Naïve Bayes was chosen as a representative model. The AUC of this model for detecting HPV+OPSCC at diagnosis was 1.0 (95% CI 0.98-1.0) (Figure 1D).

**Figure 1.**
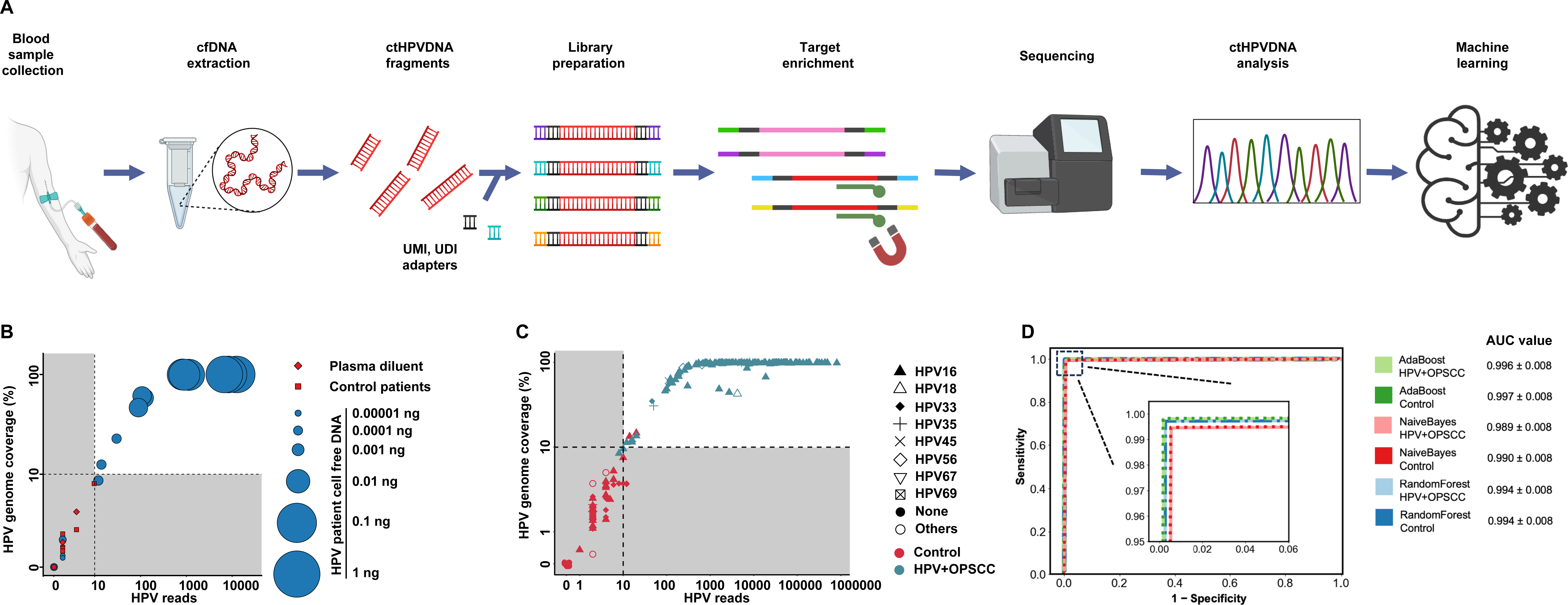
HPV-DeepSeek workflow, training and diagnostic performance. **A**. Schematic representation of HPV-DeepSeek workflow. **B**. Determination of HPV-DeepSeek threshold using serial dilutions of HPV+OPSCC patient cfDNA into control cfDNA, performed in triplicate. Circle size represents the quantity of HPV+OPSCC patient cfDNA input. Threshold was set at 10 unique HPV genome reads and 10% HPV genome coverage. Samples below the test threshold and considered negative are in the gray shaded region. White shaded region contains positive samples. The x-axis represents the number of HPV reads and the y-axis represents HPV genome coverage as percent of total genome. **C**. HPV-DeepSeek diagnostic performance in 153 HPV+OPSCC cases and 153 population-level controls applying the pre-determined thresholds. HPV+OPSCC cases are green, controls are red. HPV genotypes are represented in different shapes. Sensitivity 98.7%, Specificity 98.7%. **D**. Receiver operating curves for three machine learning models for classifying HPV+OPSCC vs. no HPV+OPSCC. The AUC values are shown for all three models based on risk scores on the test cohort with 95% confidence intervals (blue, RandomForest; green, AdaBoost; red, NaiveBayes).

### Screening detection of HPV+OPSCC

28 patients who contributed a blood sample to the MassGeneralBrigham biobank more than one year before diagnosis of HPV+OPSCC and 1:1 age and gender-matched controls were identified by database search, and cfDNA was extracted from their banked plasma samples (Table 1, Figure 2A). cfDNA from cases and controls underwent library preparation and molecular barcoding with HPV-DeepSeek and were loaded into a single next generation sequencing run. Using the prespecified cut-offs (Figure 1B, C) 22/28 (79%) of cases and 0/28 controls screened positive for HPV+OPSCC (Figure 2B). The maximum lead time from a positive screening sample to clinical cancer diagnosis was 7.8 years. The screening detection rate was 100% within four years of diagnosis. Samples that were collected closest to diagnosis generally were the furthest from the test threshold and samples collected further from diagnosis were closer to the test threshold, or negative, as expected (Figure 2C).

**Figure 2.**
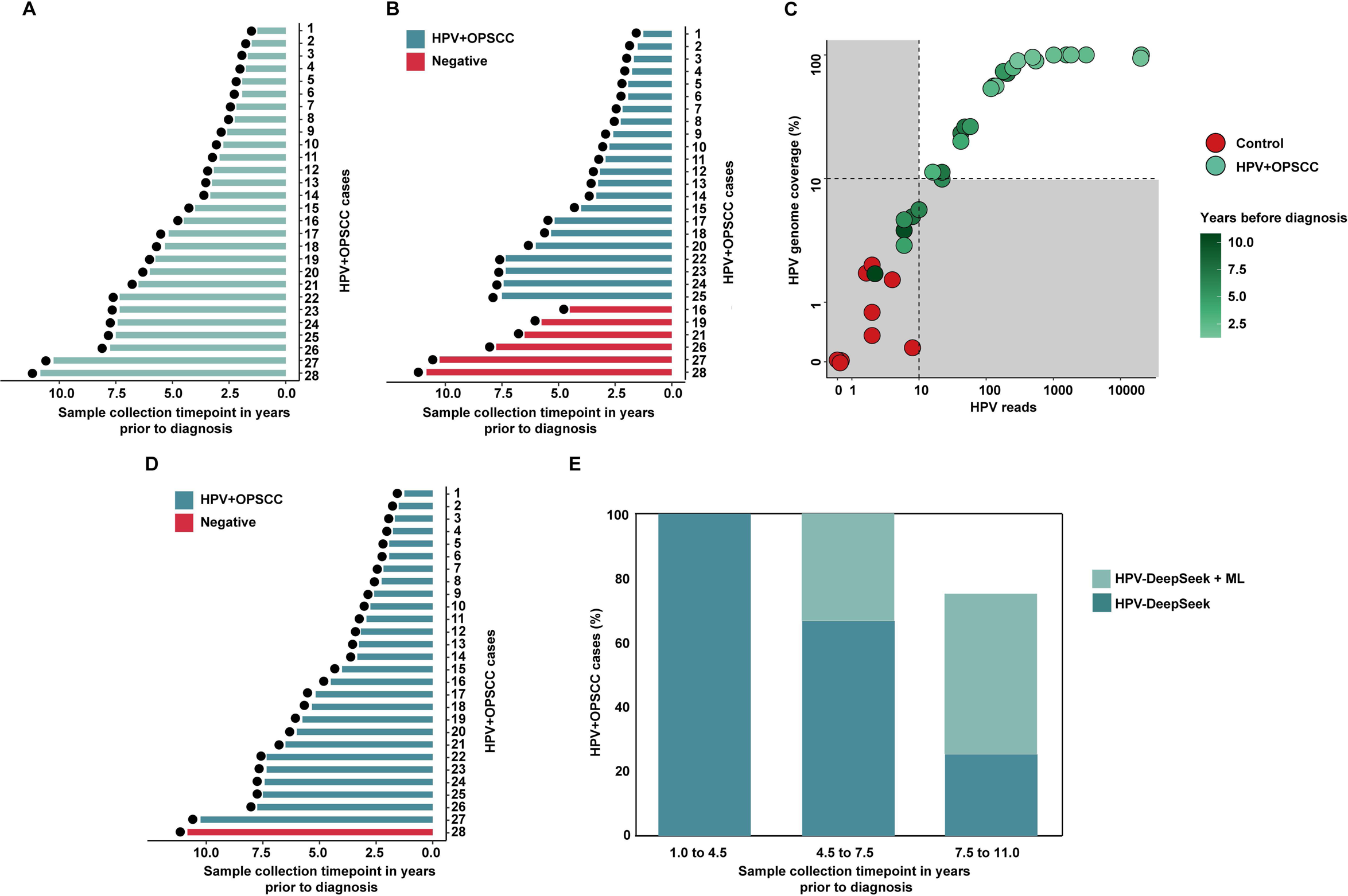
Screening detection of HPV+OPSCC. **A**. Histogram demonstrating blood sample collection timepoint in years prior to diagnosis for 28 HPV+OPSCC patients. **B**. Histogram showing results of HPV-DeepSeek in 28 HPV+OPSCC patient samples demonstrating 22/28 (79%) samples screening positive. Samples in green are positive for HPV+OPSCC based on the pre-set cutoffs. Samples in red are negative. **C**. HPV-DeepSeek results of 56 samples (28 HPV+OPSCC, green color; 28 age- and gender-matched controls, red color). The colors of green correspond to the time of collection, with light green representing samples collected closer to diagnosis and the dark green representing samples collected further from diagnosis. Lighter-green samples tend to have higher numbers of reads and higher coverage and darker-green samples tend to have lower numbers of reads and lower coverage supporting increasing ease of detection closer to time of diagnosis. **D**. Histogram showing results of HPV-DeepSeek with machine learning demonstrating 27/28 (96%) samples screening positive. **E**. Bar graph showing the comparison between HPV-DeepSeek and HPV-DeepSeek with machine learning. Data is divided in tertiles based on collection time point. The deep green color represents the percentage of cases determined positive by HPV-DeepSeek. The light green color represents the cases detected with the addition of machine learning, showing improved performance with machine learning for samples further from the time of diagnosis.

We next applied the finalized machine learning model, finding improved classification accuracy (pre-specified ctHPVDNA cutoff accuracy 0.79; Naïve Bayes accuracy 0.96, 95% CI 0.96-0.96) (Figure 2D, Supplementary Table S4, Supplementary Figure S1). The machine learning model demonstrated a maximum lead time from a positive screening sample to clinical cancer diagnosis of 10.3 years. The screening detection rate was 100% within 10 years of diagnosis. The only case not identified as HPV+OPSCC was that with the longest lead time, nearly 11 years. Machine learning improved the classification of the samples further from the time of diagnosis (Figure 2E).

### Orthogonal validation of cancer detection in the blood

Plasma-based PIK3CA gene mutations, viral integration events, and HPV serology were used to orthogonally validate detection of cancer in the blood. PIK3CA is the most mutated gene in HPV+OPSCC. 1/28 patients had pathogenic PIK3CA mutations detected by HPV-DeepSeek (Figure 3A, Supplementary Figure S2). Viral genome integration into the human genome is a hallmark of HPV-associated cancers. 7/28 patients had viral integration detected by HPV-DeepSeek (Figure 3A, B). HPV oncoprotein antibodies have high sensitivity and specificity for HPV+OPSCC at the time of diagnosis. Utilizing a 5- genotype multiplexed serology assay, 26/28 cases were tested for antibodies with 20/26 (77%) of cases having detectable antibodies and 17/26 (65%) of cases having both ctHPVDNA and HPV antibodies (Figure 3A, 3C, Supplementary Table S5). 19/28 cases had at least two cancer diagnosis-supporting features detected in the plasma (Figure 3D).

**Figure 3.**
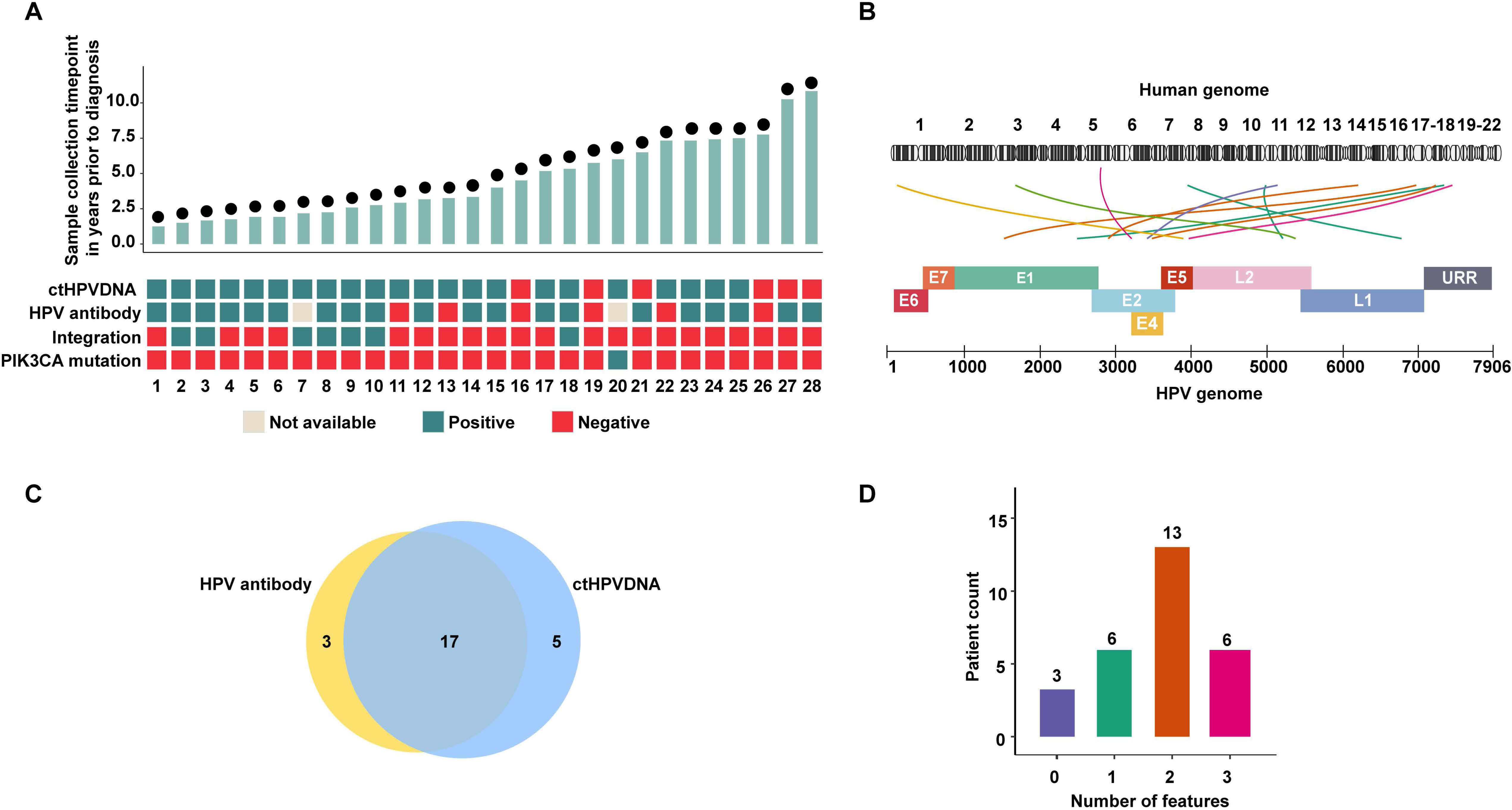
Orthogonal validation of cancer detection in the blood. **A**. Histogram representing the 28 HPV+OPSCC samples ordered by sample collection time in years prior to diagnosis. Below, features supportive of cancer detection. Samples positive for a given feature are in green, samples negative for a feature are in red, and untested samples are in beige. **B**. HPV-human genome integration events detected in seven patients. **C**. Venn diagram for HPV serology and ctHPVDNA results showing 20/26 patients had HPV oncoprotein antibodies detected of which 17 also had ctHPVDNA detected**. D**. Bar plot indicating the number of samples with 0, 1, 2 or 3 cancer diagnosis-supporting features present. 25/28 cases had at least one cancer diagnosis supporting feature and 19/28 had at least two features.

### Viral genome molecular fingerprinting

To ensure contamination could not account for screening detection we utilized viral genome molecular fingerprinting between and within cases. 6/22 cases that screened positive displayed unique HPV genotypes or sub-lineages (Figure 4A). As expected from epidemiologic studies, some viral genomes shared the same HPV16 sub-lineages, with 12/22 cases belonging to the HPV16 A1 sub-lineage, 3/22 being D3 and 2/22 being A2. Within shared sub-lineage samples, we evaluated SNVs within each viral genome demonstrating that 13/17 viruses displayed unique SNV fingerprints while the remaining four lacked SNVs, precluding assignment as unique viruses (Figure 4A, Supplementary Figure S3).

**Figure 4.**
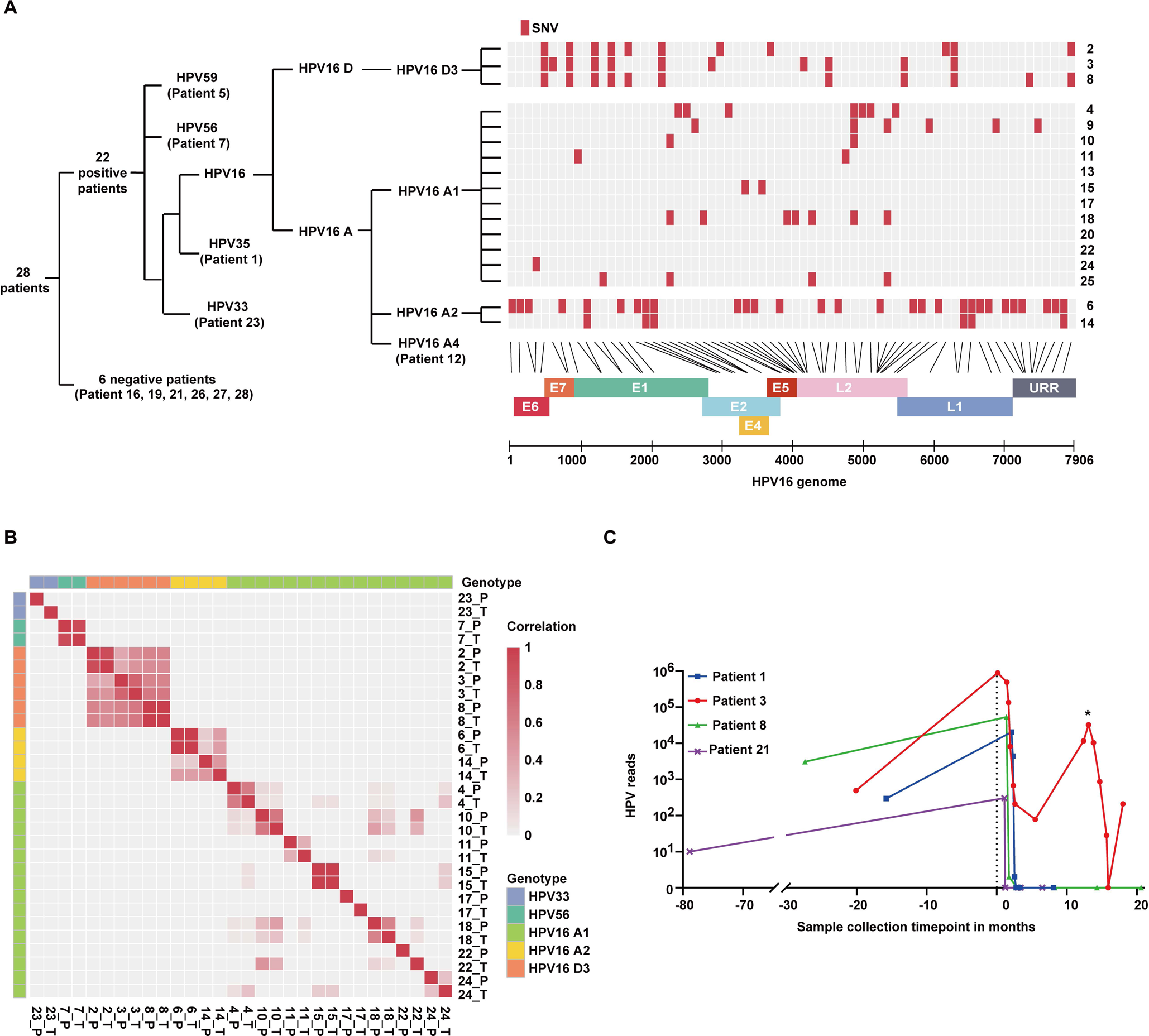
Viral genome molecular fingerprinting and longitudinal cancer monitoring. **A**. Phylogenetic tree of the 28 HPV+OPSCC samples based on the genotype, lineage and sublineage detected in plasma (left) and molecular fingerprinting heatmap (right) for samples sharing the same HPV16 sublineage. SNVs for each sample are shown in red, demonstrating that each virus is unique. **B**. Pairwise correlation heatmap for the 15 samples that had FFPE tumor tissue blocks from diagnosis available, showing that plasma-tissue pairs correlate within a pair and not across pairs. The highest correlation of 1 is represented in red while no correlation or 0 is represented in white. **C**. Longitudinal monitoring of four HPV+OPSCC patients from screening time point to diagnosis, treatment and post-treatment monitoring. Vertical dotted line represents time of diagnosis. Patients 8 and 21 were treated with surgery. Patients 1 and 3 were treated with chemoradiotherapy. In patients 1, 8, and 21 ctHPVDNA is cleared following treatment and remained zero during monitoring. In patient 3, ctHPVDNA was detected 20 months before diagnosis. ctHPVDNA levels were monitored weekly during chemoradiotherapy treatment with decreasing levels, but no clearance. Following conclusion of treatment, ctHPVDNA levels began increasing. The patient was then found to have a second primary HPV malignancy (asterix) for which they underwent surgery followed by chemoradiotherapy. ctHPVDNA cleared after this treatment but re-elevated, indicating recurrence, which was detected by cross-sectional imaging two months later.

15/28 patients were treated for their eventual cancer diagnosis within our healthcare system and had FFPE tumor tissue blocks which could be retrieved from diagnostic biopsy or surgery. DNA from tumor tissue was subject to HPV-DeepSeek and viral genome fingerprinting was performed to verify the same viral genome in both pre-diagnostic plasma and diagnostic tumor tissue. All plasma-tissue pairs shared the same genome, with matching only to the pair-mate, and to no other viral genome, confirming that the same virus was present from screening detection to diagnosis (Figure 4B, Supplementary Figure S4).

### Longitudinal monitoring

7/28 patients had been prospectively enrolled into ctHPVDNA studies within our lab at the time of cancer diagnosis unrelated to their biobank sample collection (Supplementary Figure S5). Four of these patients had samples collected at the time of clinical cancer diagnosis and into treatment monitoring affording a continuous global view from cancer screening detection through post-treatment monitoring (Figure 4C). In one of these patients, ctHPVDNA persisted across treatments, predicting recurrence.

## Discussion

Using HPV+OPSCC as a model, we demonstrate accurate detection of HPV-associated cancers with 100% sensitivity and specificity within four years of clinical cancer diagnosis and a maximum lead time from detection to clinical diagnosis of 7.8 years. The application of machine learning extended detection up to ten years prior to clinical diagnosis. These results were validated by examining orthogonal blood-based features that support cancer detection such as PIK3CA gene mutations, viral integration events and HPV oncoprotein antibodies. Contamination was ruled out by demonstrating that each patient harbored a unique viral genome. In patients with tumor tissue available from diagnosis, the same viral genome was seen in blood screening detection and tissue diagnosis. These results highlight the enormous potential of ctDNA detection approaches for cancers that currently lack screening tests, and further demonstrate some of the most significant lead times from molecular cancer detection to clinical diagnosis reported.

Virally induced cancers have been a long-standing model system for early detection studies as they afford a unique ctDNA target, simplifying the search for these rare cancer fragments in the blood, and because they induce unique immunologic responses. In EBV-associated nasopharyngeal carcinoma, Lo and colleagues have demonstrated the ability to detect asymptomatic cancers using circulating tumor EBV DNA, and that this detection leads to diagnosis of earlier stage disease and improved survival^26^. Serologic markers of EBV infection have also been shown to be an effective screening approach for EBV-associated nasopharyngeal carcinoma in select populations^27^. Importantly, cancers induced by infectious agents account for 15-20% of all malignancies worldwide. HPV is the cause of 5% of all malignancies, accounting for 684,000 new cancer diagnoses per year^28–30^. While Pap smear screening has led to a significant decline in cervical cancer incidence in high-income countries, cervical cancer remains the second leading cause of cancer death for women worldwide, and no population-level screening tests are utilized for the remaining five HPV-associated cancers, including HPV+OPSCC, the most common HPV-associated malignancy in the United States.

HPV+OPSCCs lead to measurable immunologic response through HPV oncoprotein antibodies, a potential screening biomarker. However, the long and variable time between seroconversion and HPV+OPSCC development, and the non-linear nature of the biomarker, means HPV oncoprotein seropositivity has limited clinical utility. Further, the performance characteristics of HPV oncoprotein seropositivity in anogenital HPV-associated cancers are significantly worse^31^. ctHPVDNA detection has emerged as a promising biomarker for diagnosing and monitoring HPV+OPSCCs, which is being widely applied across multiple clinical contexts^20,32–34^. The potential of ctHPVDNA as a screening tool for HPV- associated cancers has been hampered by limitations in the sensitivity of available assays for ultra-low ctDNA levels expected in a screening setting. However, HPV+OPSCC presents an ideal scenario for studying blood-based early cancer detection considering the long natural history of HPV infection in the oropharynx, and the presumed development of HPV+OPSCC years or even decades prior to clinical diagnosis, supported by HPV oncoprotein studies and genomic modeling^15–19^. The findings of this study, demonstrating remarkable lead times from molecular detection to clinical diagnosis, fit with this presumed natural history.

We evaluated the relationship between HPV serology and ctHPVDNA and found that ctHPVDNA had at least equivalent sensitivity to HPV serology overall (ctHPVDNA 79% vs Serology 77%) with a trend for improved sensitivity of ctHPVDNA closer to diagnosis (ctHPVDNA 100% vs Serology 86% within four years of diagnosis). ctHPVDNA has several additional advantages over serologic testing, including that approaches for detecting ctHPVDNA are more widely available and reproducible, and that ctHPVDNA is a dynamic biomarker that reflects disease burden and thus can be trended across time and clinical contexts, as evidenced by the longitudinal monitoring presented.

Our findings raise practical and ethical questions, including how practitioners or patients should act on a positive ctHPVDNA screening test, considering these molecularly-detected cancers are likely not clinically apparent. While our study demonstrates the feasibility of blood-based detection of HPV- associated cancers years prior to clinical diagnosis, it does not attempt to determine whether early detection would benefit patients. While it is presumed that earlier detection of asymptomatic cancers would be beneficial, several questions must be addressed to prove this, including whether earlier detection leads to improved survival, lower cost, and decreased treatment morbidity, and whether patient anxiety and the recently described “cancer patient in waiting” scenario can be adequately managed to attain overall improvement in quality of life^35^. Long-term, we envision early detection of HPV-associated malignancies leading to treatment with tolerable, low-morbidity approaches such as personalized cancer vaccines, with the presumption that early cancers are more therapeutically tractable and thus some could be ablated before ever becoming clinically apparent, while others may not respond and would need to be monitored molecularly until they became clinically apparent. The development of screening methods as demonstrated here must therefore occur in concert with the development of treatment and monitoring plans and approaches to address the psychosocial dimensions of such scenarios. Further, the appropriate selection of patients and timing for screening requires careful consideration and study. Multiple potential approaches have been proposed, including serial screening of those at highest risk for HPV-associated malignancies, such as individuals with HIV, single screening timepoints for moderate risk inviduals, such as men at age 50, or population-level application through the inclusion of ctHPVDNA as one marker in an MCED^36^. The utility, cost, and population-level diagnostic performance of these and other screening approaches will require rigorous prospective study.

In conclusion, using plasma samples from asymptomatic individuals who went on to develop HPV+OPSCC and healthy population-level control patient samples, we demonstrate sensitive and specific blood-based detection of HPV-associated cancers extending to a decade prior to clinical cancer diagnosis. These results highlight the enormous potential of ctDNA screening approaches and raise the opportunity for a blood-based screening test for 5% of all cancers worldwide.

## Data Availability

All data produced in the present work are contained in the manuscript

