## Supplemental for "Blood-based screening for HPV-associated cancers"

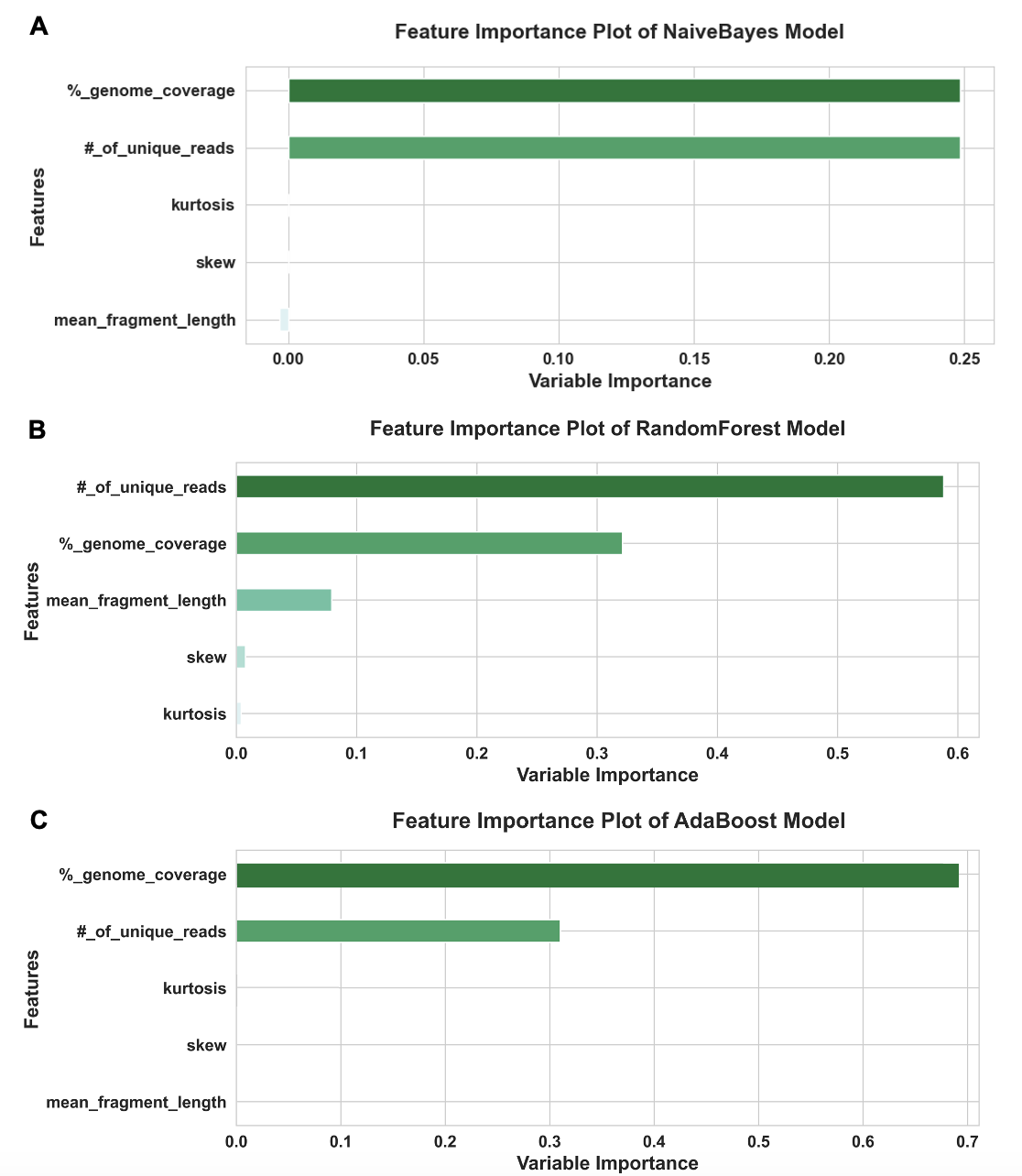
**Supplementary Figures**

**Supplementary Figure S1. Comparative Visualization of Model Feature Importance**

Bar plots, each corresponding to a different machine learning model: NaiveBayes (top), RandomForest (middle), and AdaBoost (bottom). Each plot illustrates the relative importance of five distinct features used by the respective models to perform classification tasks. The features are listed on the y-axis in descending order of importance. The x-axis represents the variable importance, indicating how much each feature contributes to the model's predictive power. This comparative analysis aids in understanding the behavior of different models and the significance of each feature in the classification process.

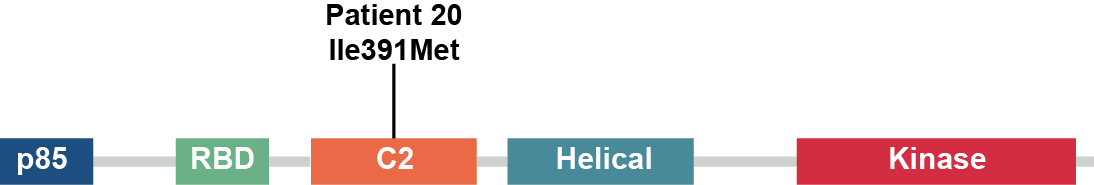

**Supplementary Figure S2. Domain-specific PIK3CA Mutation Distribution in Patient Cohort**

Lollipop plot illustrating the Ile391Met PIK3CA mutation in patient number 20. Different regions of the PIK3CA protein are marked in different colors representing p85 (p85-binding domain), RBD (Ras-binding domain), C2 (protein kinase C conserved domain 2), Helical, and Kinase domains.

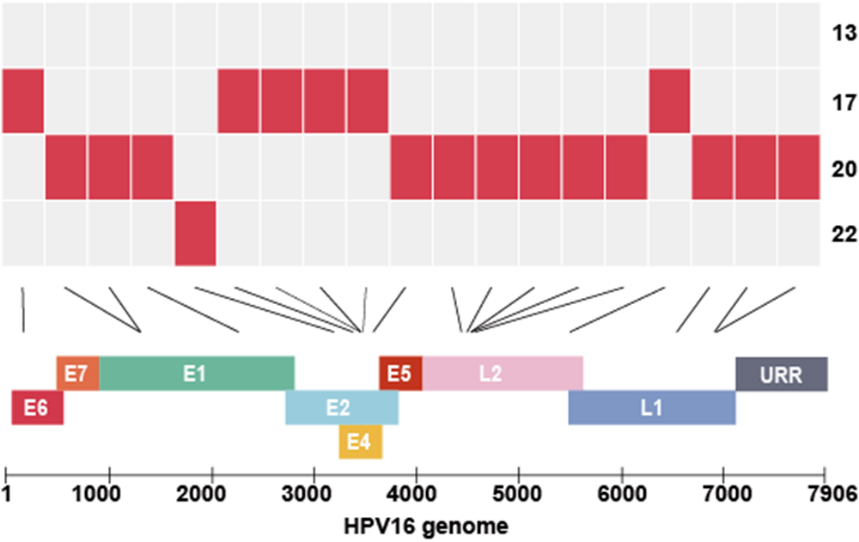

**Supplementary Figure S3. Mutation Profile of HPV16 Genomes Across Different Patients**

Mutation analysis of HPV16 genomes from four patients (13, 17, 20, and 22). The top panel displays the distribution of mutations across the viral genome for each patient, represented by red blocks. The analysis was carried out without filtering threshold (AF> 0.5 & Depth≥4 reads) for the identification of all single nucleotide variations (SNVs), providing a complete molecular fingerprint for each viral sample. The bottom panel maps the HPV16 genome, with colored blocks representing different coding regions: E6, E7, E1, E2, E4, E5, L2, and L1, and the untranslated region (URR). Arrows between the panels indicate the corresponding locations of mutations on the genome map. This unrestricted mutation plot aids in understanding the unique mutational landscape of HPV in each sample.

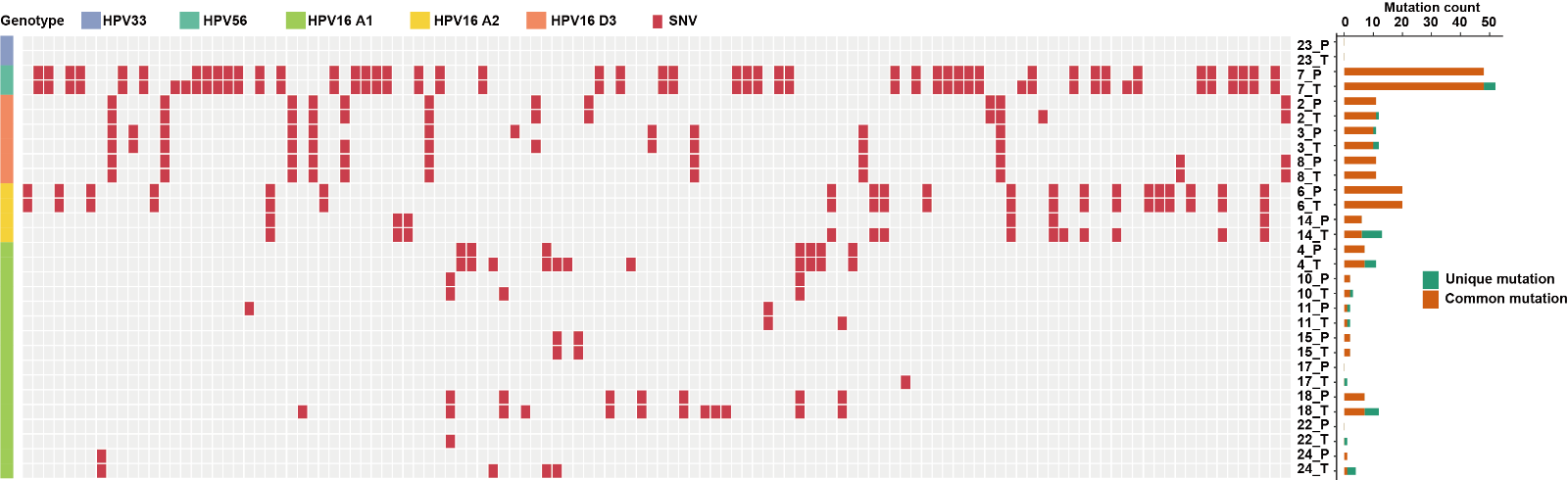

**Supplementary Figure S4. Analysis of Single Nucleotide Variants (SNVs) in Matched Blood and Tumor Samples**

Heatmap demonstrating the analysis of Single Nucleotide Variants (SNVs) in matched blood and tumor samples showing the HPV genome similarity between plasma and tumor tissues. Each row corresponds to an individual patient sample, while each column represents a specific mutation. The presence of a mutation is indicated by red. The samples are labeled as P (Plasma) and T (Tumor) and the corresponding patient number. The bar graph on the right side represents the mutation count of each viral genome. The mutations shared between paired plasma and tumor are shaded in orange while the green bar represents the mutations that appear only in one sample.

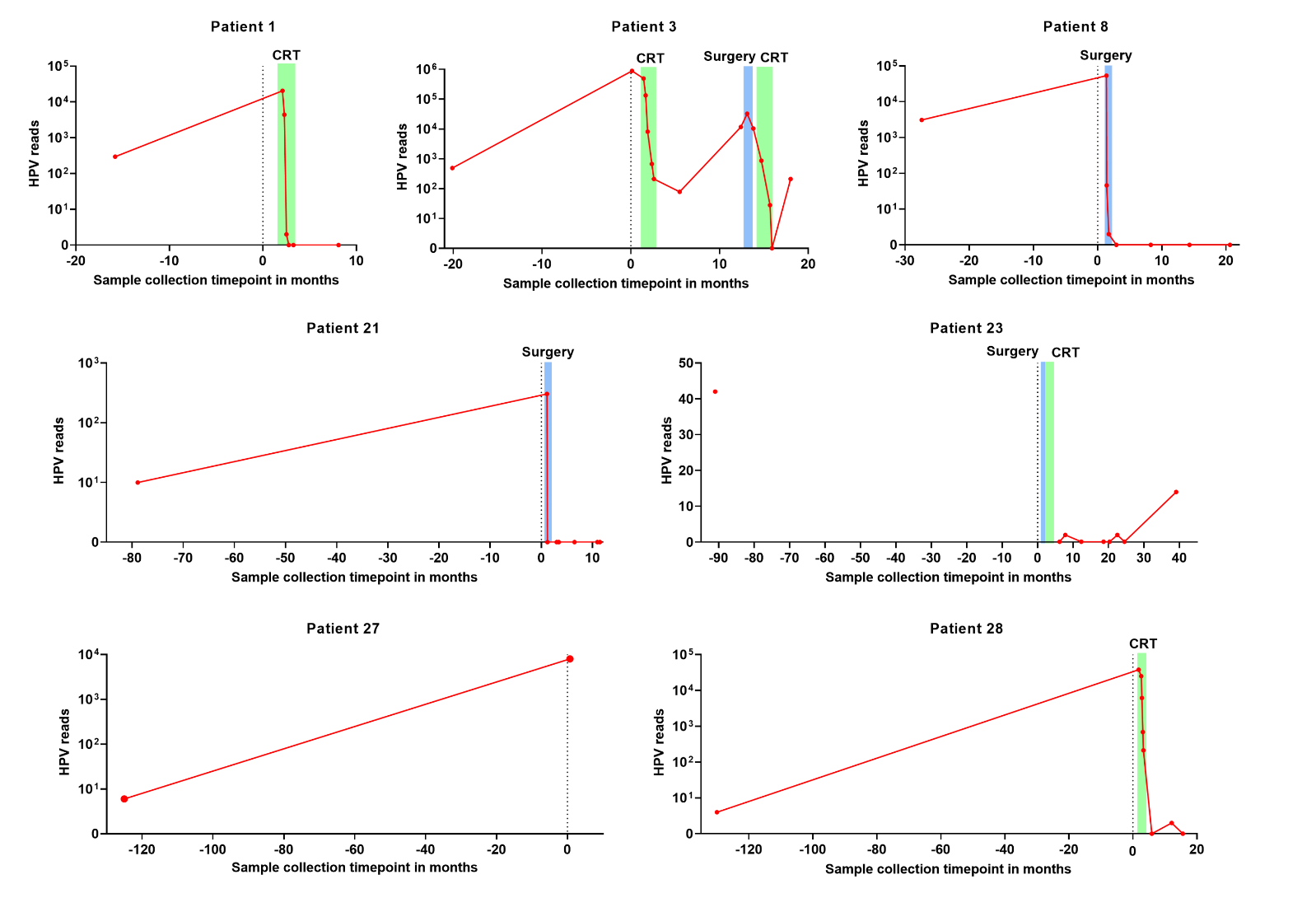

**Supplementary Figure S5. Longitudinal ctHPVDNA Monitoring**

Changes in circulating tumor HPV DNA (ctHPVDNA) over time for seven patients. Each plot represents one patient's ctHPVDNA trajectory as a function of time, with the sample collection timepoint in months on the x-axis and the ctHPVDNA loads on the y-axis, plotted on a logarithmic scale. The vertical dashed lines represent the date of diagnosis. The data points are plotted as red dots, and the connecting lines indicate the trend over the observed period.

*Patient 1*. ctHPVDNA was detected 16 months before diagnosis. ctHPVDNA levels were monitored weekly during chemoradiotherapy (CRT) treatment with clearance at the end of treatment and remaining zero during monitoring. *Patient 3*. ctHPVDNA was detected 20 months before diagnosis. ctHPVDNA levels were monitored weekly during CRT treatment with decreasing levels but no clearance. Following conclusion of treatment, ctHPVDNA levels began increasing. The patient was then found to have a second primary HPV malignancy for which they underwent surgery followed by CRT. ctHPVDNA cleared after this treatment but re-elevated, indicating recurrence, which was detected by cross-sectional imaging two months later. *Patient 8*. ctHPVDNA was detected 27 months before diagnosis. ctHPVDNA rapidly cleared following surgery and remained zero during monitoring. *Patient 21*. ctHPVDNA was not detectable 79 months before diagnosis but was detected at the time of clinical diagnosis. The patient was treated with surgery. ctHPVDNA cleared rapidly with surgery and remained zero during monitoring. *Patient 23*. ctHPVDNA was detected 91 months before diagnosis. A sample at diagnosis was not available. ctHPVDNA was negative after treatment with surgery and CRT but subsequent monitoring showed an increase, indicating a recurrence. *Patient 27*. ctHPVDNA was not detectable 125 months before diagnosis, but was detected at the time of diagnosis. *Patient 28*. ctHPVDNA was not detected 130 months before diagnosis but was detected at diagnosis. The patient underwent weekly ctHPVDNA monitoring during CRT with clearance at the end of treatment and remained zero during monitoring. This figure emphasizes the potential of an integrated screening, diagnosis, and monitoring biomarker.

**Supplementary Tables**

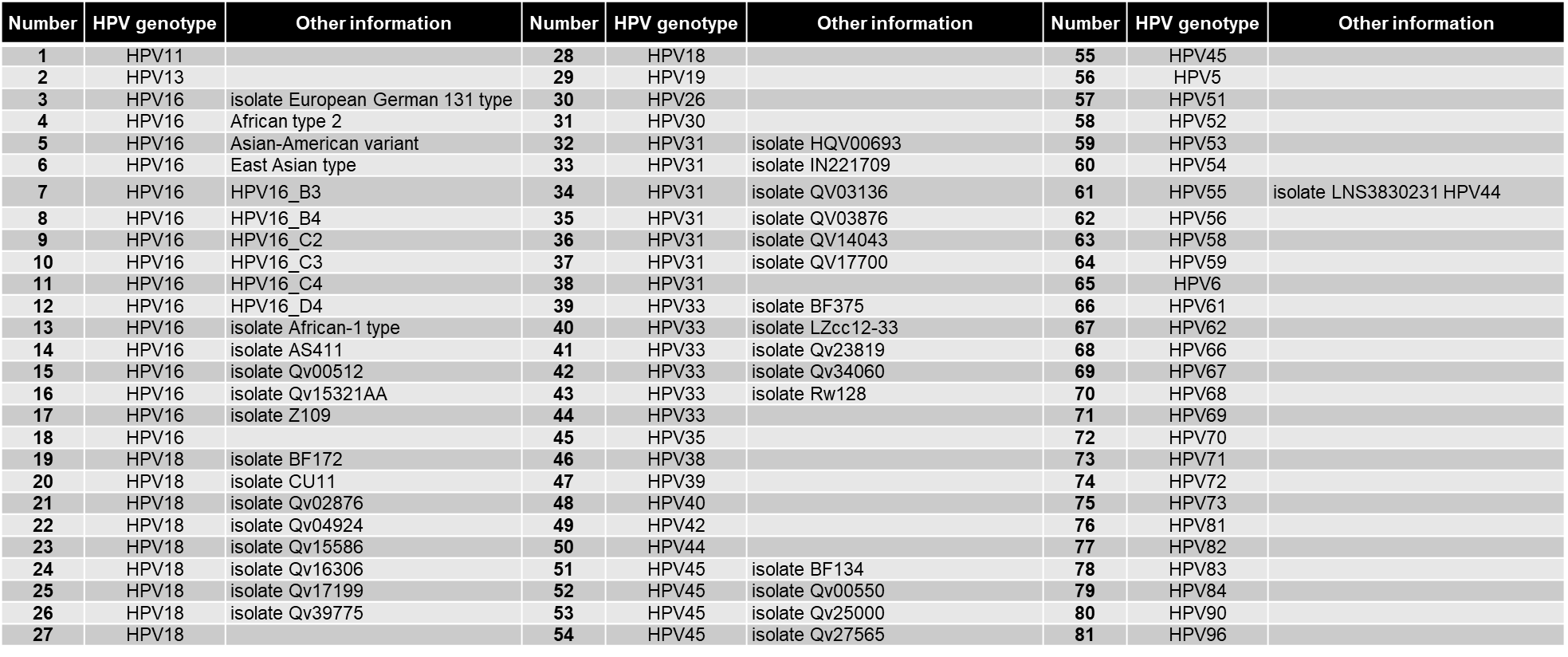

**Supplementary Table S1. Inventory of HPV Genotyping Probes Used in Capture Library**

List of HPV genotypes targeted by probes in the genomic capture library. Each entry in the table corresponds to a unique HPV genotype or sublineage. Additional details for certain genotypes are provided, which include isolate names, variant designations, or subtype information relevant for differential genomic analysis.

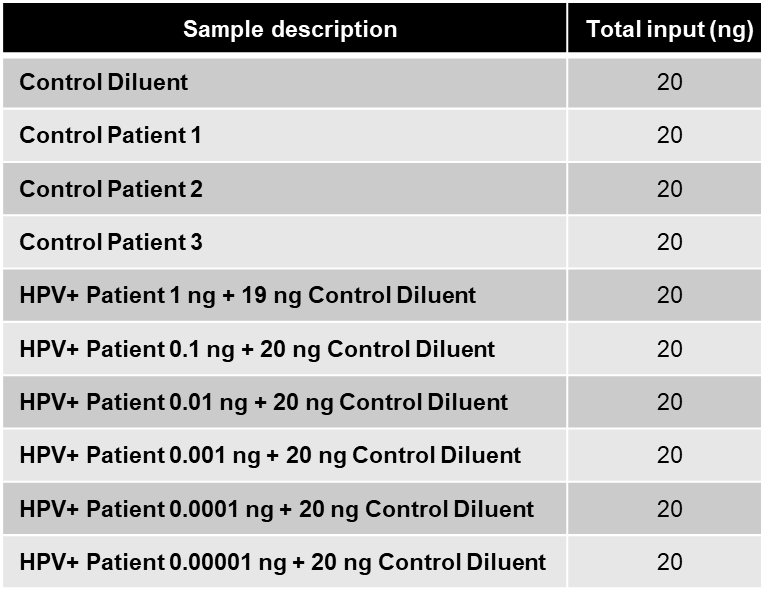

**Supplementary Table S2. Inputs For Limit of Detection Determination**

Total input in nanograms of DNA from HPV patient samples and control samples used to determine the limit of detection for HPV-DeepSeek. Each row indicates the combination of patient sample DNA and control diluent used to achieve a total input of 20 ng per reaction. Control samples include a pure control diluent and three different patient controls to establish baseline readings. The HPV patient samples are serially diluted with control diluent to demonstrate the sensitivity of the assay across a range of HPV DNA concentrations from 1 ng to 0.00001 ng.

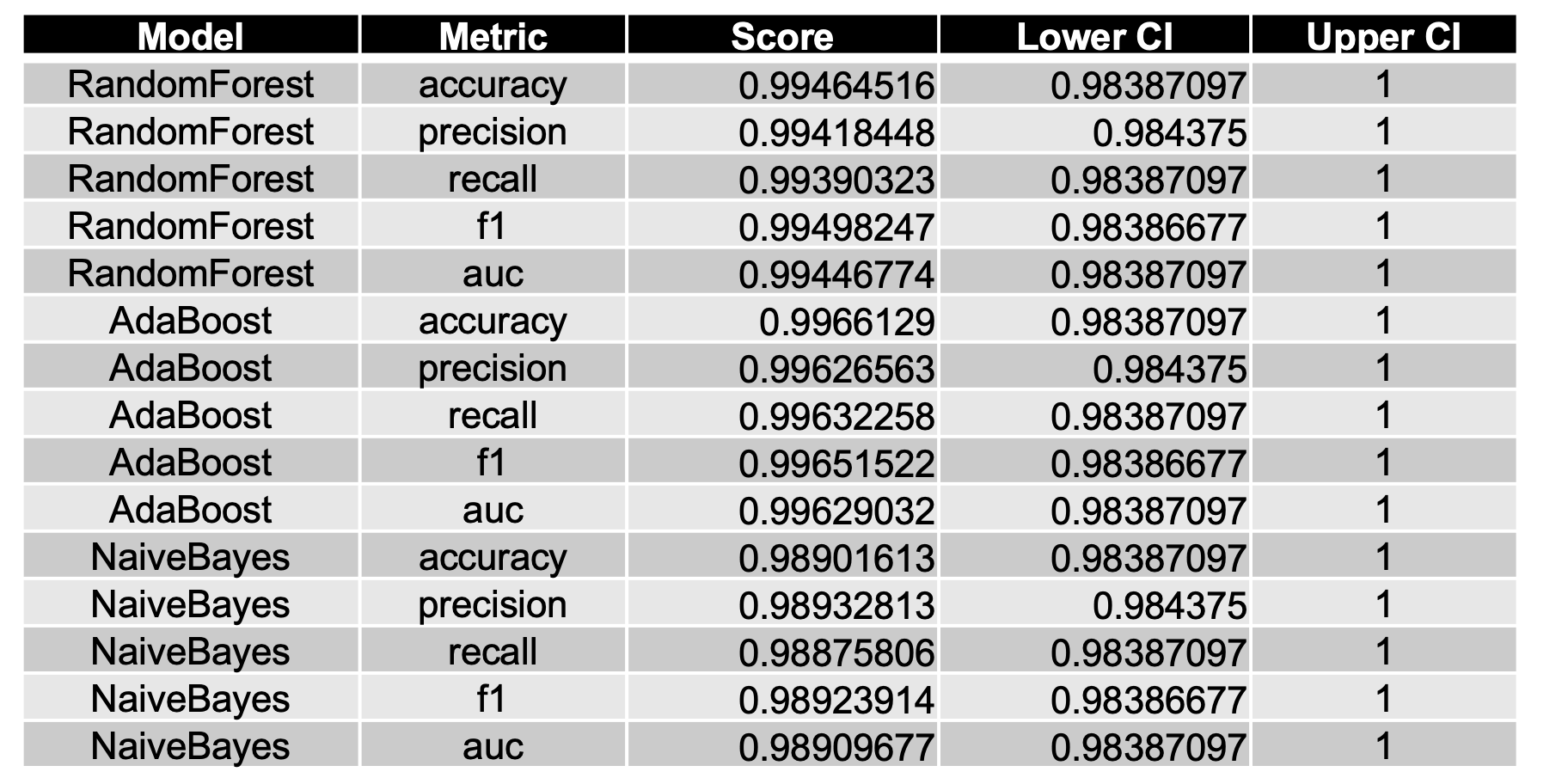

**Supplementary Table S3. Comparative Performance Metrics of Machine Learning Models on Test Set**

This table presents the performance evaluation of three bootstrapped machine learning models – NaiveBayes, RandomForest, and AdaBoost – using five metrics on the test dataset. The metrics used for assessment are accuracy, precision, recall, f1 score, and area under the receiver operating characteristic curve (AUC). Each model’s performance is quantified by a score, alongside the lower and upper bounds of the 95% confidence interval (CI) for each metric. The parameters for bootstrap resampling were 1,000 repetitions from a sample of 50% of the original dataset.

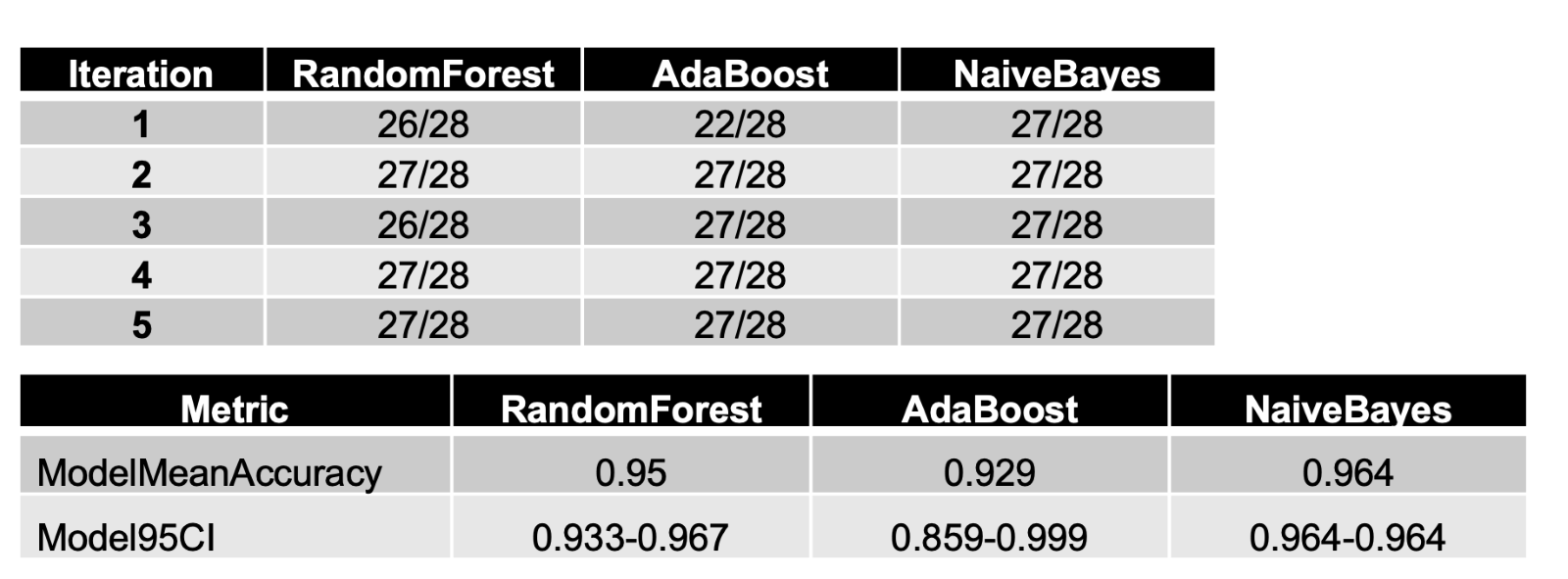

**Supplementary Table S4. Iterative Performance and Confidence Intervals of Machine Learning Models on Validation Set**

Analysis of the performance of three machine learning models – NaiveBayes, RandomForest, and AdaBoost on the validation dataset. Performance is evaluated over five iterations, with each iteration's result expressed as a fraction of successful predictions out of 28 cases. The upper table shows the raw performance data for each iteration, while the lower provides summary metrics. This includes the mean accuracy of the model across all iterations and the 95% confidence interval (CI) for the model accuracy.

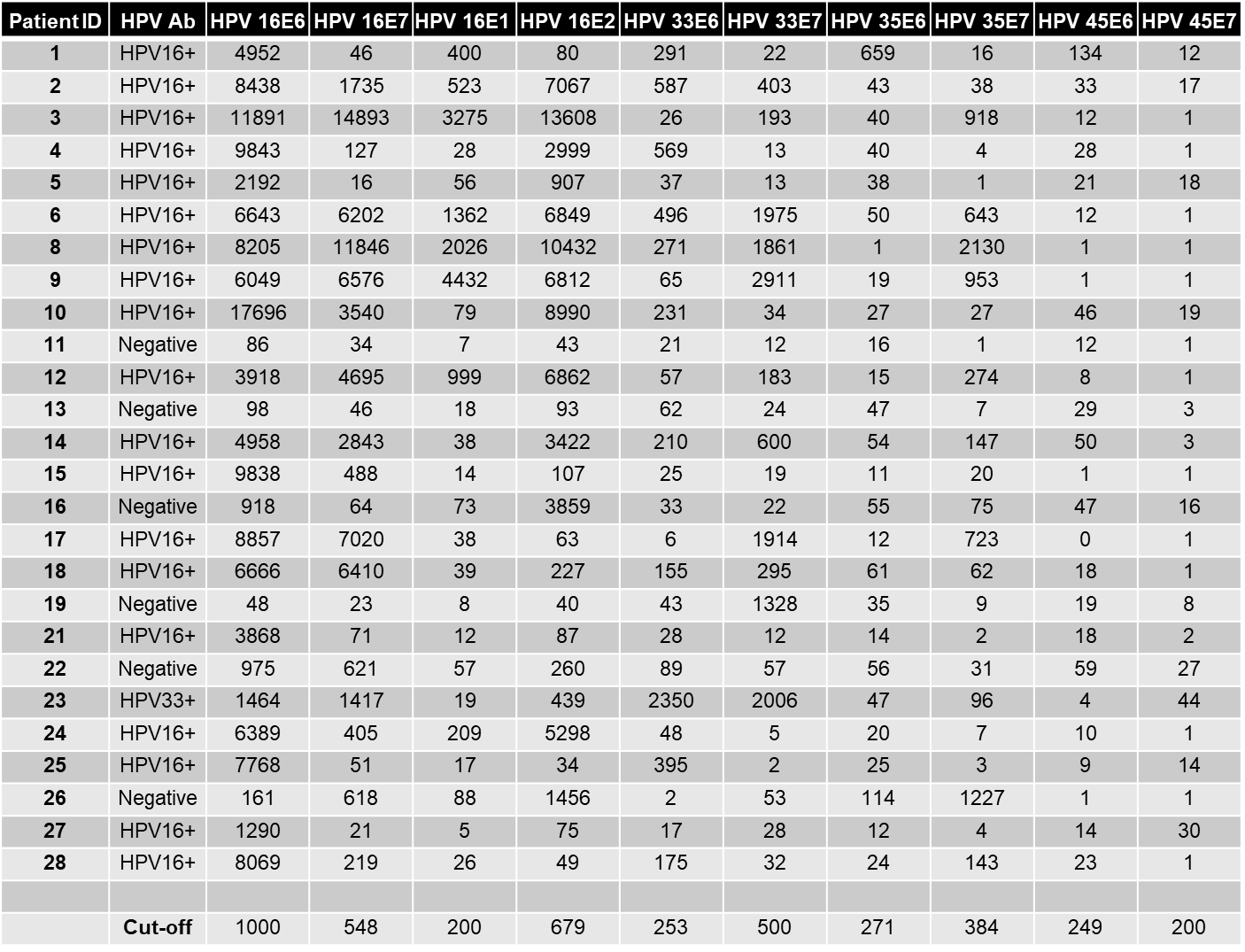

**Supplementary Table S5. HPV Multiplexed Serology Assay**

This table presents the findings of a multiplex serology assay for HPV, conducted on a sample set of 28 cases. The serological responses are quantified as median fluorescence intensities (MFIs) for multiple HPV genotypes and their respective antigens. The assay targets antibodies against multiple epitopes of HPV16, including E6, E7, E1, and E2, as well as other high-risk HPV types 33, 35, and 45, using their E6 and E7 antigens. A case is classified as HPV16 antibody positive if the MFI for HPV16 E6 alone is above the cut-off, or if at least three out of four antigens (E1, E2, E6, E7) related to HPV16 are above their respective cut-offs. In this instance, the E6 cut-off is specifically adjusted to 484 MFI. For non-HPV16 types (HPV33, HPV35, and HPV45), positivity requires both E6 and E7 MFIs to exceed their respective cut-off values. Some cases exhibited positivity for multiple HPV genotypes, which could indicate cross-reactivity in the assay. Where applicable, type specific E6 ratios were calculated for HPV16 and HPV33. Finding HPV16 E6 MFIs were considerably higher than the HPV33 E6 MFIs, these cases were classified as HPV16 positive by serology. Assay results are arranged in rows for each patient, with the patient ID listed in the first column, and subsequent columns indicating the MFI readings for each antigen. The row at the bottom of the table specifies the cut-off MFI values used to determine seropositivity.
